# Estimating heterogeneity of treatment effect in psychiatric clinical trials

**DOI:** 10.1101/2024.04.23.24306211

**Authors:** Joshua S. Siegel, Jinglin Zhong, Sasagu Tomioka, Ajay Ogirala, Stephen V. Faraone, Steven T. Szabo, Kenneth S. Koblan, Seth C. Hopkins

## Abstract

Currently, placebo-controlled clinical trials report mean change and effect sizes, which masks information about heterogeneity of treatment effects (HTE). Here, we present a method to estimate HTE and evaluate the null hypothesis (H_0_) that a drug has equal benefit for all participants (HTE=0). We developed measure termed ‘estimated heterogeneity of treatment effect’ or ***eHTE***, which estimates variability in drug response by comparing distributions between study arms. This approach was tested across numerous large placebo-controlled clinical trials. In contrast with variance-based methods which have not identified heterogeneity in psychiatric trials, reproducible instances of treatment heterogeneity were found. For example, heterogeneous response was found in a trial of venlafaxine for depression (p_eHTE_=0.034), and two trials of dasotraline for binge eating disorder (Phase 2, p_eHTE_=0.002; Phase 3, 4mg p_eHTE_=0.011; Phase 3, 6mg p_eHTE_=0.003). Significant response heterogeneity was detected in other datasets as well, often despite no difference in variance between placebo and drug arms. The implications of eHTE as a clinical trial outcomes independent from central tendency of the group is considered and the important of the eHTE method and results for drug developers, providers, and patients is discussed.

## 1 Introduction

You have heard some variation of the refrain – “Evidence-based medicine is derived from group averages, yet medical decisions are made by and for individuals” (Kent, 2023). The barrier to more personalized medicine: the largest and most expensive clinical trials conducted are reported with group means and effect sizes. The clinical problem of individualized medicine is related to the statistical problem of the heterogeneity of treatment effects (HTE), also known as treatment heterogeneity (Longford, 1999). This concept is crucial in clinical research and healthcare because it acknowledges that the efficacy or safety of treatments can differ among patients due to a variety of factors, such as age, gender, genetic makeup, disease severity, coexisting conditions, and environmental influences (Miller & Raison, 2023).

Nowhere is the problem with this system better illustrated than in clinical studies on antidepressants. Selective serotonin reuptake inhibitors are the first line treatment for depression and are among the most prescribed medications. Yet in large meta-analyses, the average treatment effect of antidepressants is found to be about two points on the HAMD-17 scale (Cipriani et al., 2018). This is below the assumed clinically relevant effect (3-7 points) (Moncrieff & Kirsch, 2015). Despite the small average treatment effect of antidepressants, it is commonly assumed that subpopulations of patients exist that have a clinically relevant benefit (Fava, 2015). However, heterogeneity of treatment effect, while widely believed and intuitively plausible, has not been shown to exist. On the contrary, because the variability in antidepressant and placebo arms are roughly equal, some have inferred that response to antidepressants is not heterogenous (Maslej et al., 2021; Volkmann et al., 2020).

HTE is the magnitude of variation of individual treatment effects across a population. It is sometimes regarded as dichotomous – present, or absent. If there is no interaction between treatment and individual, then there is no HTE. Conversely, HTE is present when the same treatment produces different results in different patients (Sørensen, 1996). When HTE is present, a modest benefit (such as is observed with antidepressants) can be misleading because modest average effects may reflect a mixture of substantial benefits for some, little benefit for many, and harm for a few.

Non-zero HTE is an implicit (typically untested) assumption in a broad literature of statistical approaches for clinical trial subgroup analysis (Senn, 2018). But a tool to explicitly test for treatment heterogeneity in existing clinical trials is lacking. It would provide an important guidance on when enrichment and subgroup analysis might be of benefit.

HTE is the standard deviation of the individual treatment effect (ITE) rather than the standard deviation of the outcomes in the target population. ITE is the hypothetical difference between a person’s outcome on treatment A and their outcome on treatment B (often placebo) (Kravitz et al., 2004). In most clinical trials, it is impossible to measure ITE directly. Thus, it is impossible to precisely measure heterogeneity. But, by assuming that response to active drug = placebo effect + drug effect, it becomes possible to estimate treatment heterogeneity in an adequately powered placebo-controlled trial.

If active drug response = placebo effect + treatment effect, one can test the null hypothesis (H_0_) that an effective drug homogenously shifts the distribution of placebo responses over (ITE=0). Below, we describe a simple approach which compares the distribution of responses between arms of a study (typically placebo and active treatment). We show that measuring the standard deviation of response difference between any patient on treatment and the corresponding placebo patient with the matching percentile is a close approximation of the HTE. We then use simulated and real clinical trial datasets to describe different types of heterogeneity and also document that heterogeneity is present in psychiatric clinical trials.

## 2 Methods

### 2.1 Overview

To assess and compare treatment heterogeneity in randomized controlled trials (RCTs), we employed a computational approach first and then tested the resulting framework on real participant-level RCT data (datasets in which baseline and outcomes data were available for every participant, rather than group statistics only). This approach focused on estimating the variation in treatment responses among individual participants (ITE), between placebo and active treatment groups, rather than merely comparing average responses and treatment effects.

This method was applied first to a set of “toy” cases, to provide an intuition for how our conceptual framework of heterogeneity relates to the methodological implementation, and then to multiple previously published placebo-controlled clinical trials, to explore the presence of treatment heterogeneity across multiple therapeutic areas and interventions. The outcomes of different treatment arms were compared through the computed eHTE (and associated *P*-values), enabling the identification of the presence and magnitude of heterogeneity in each trial.

### 2.2 Cumulative Response Curves

For each treatment arm, individual participants’ outcome scores (e.g., change in MADRS, in a depression trial) were used to plot cumulative response curves, which showcase the distribution of responses within each arm. Each score represents the change in the primary endpoint from the baseline, allowing visualization of the relative proportions of individuals reporting varying degrees of response.

### 2.3 eHTE - an estimate of heterogeneity of treatment effect

Conceptually, eHTE can be thought of as the variability of individual treatment effect (interaction of individual x treatment) proportional to variability in clinical response due to all other causes (variance of the placebo response). This was accomplished as follows: we first sorted participants in each arm by ascending magnitude of response on the primary outcome and plotted response across percentiles (Fig 1A). We then calculated 48 pairwise drug-placebo differences (a conservative approximation of individual treatment effect) across percentiles (ranging from 3^rd^,5^th^,7^th^ …97^th^ percentiles, to reduce the influence of outlier participants) and finally computed the standard deviation across all 48 drug-placebo differences. The resulting value was normalized by dividing by the standard deviation in placebo response. Thus, eHTE is the standard deviation of response difference across drug-placebo pairs divided by the standard deviation in placebo response.

**Figure 1.**
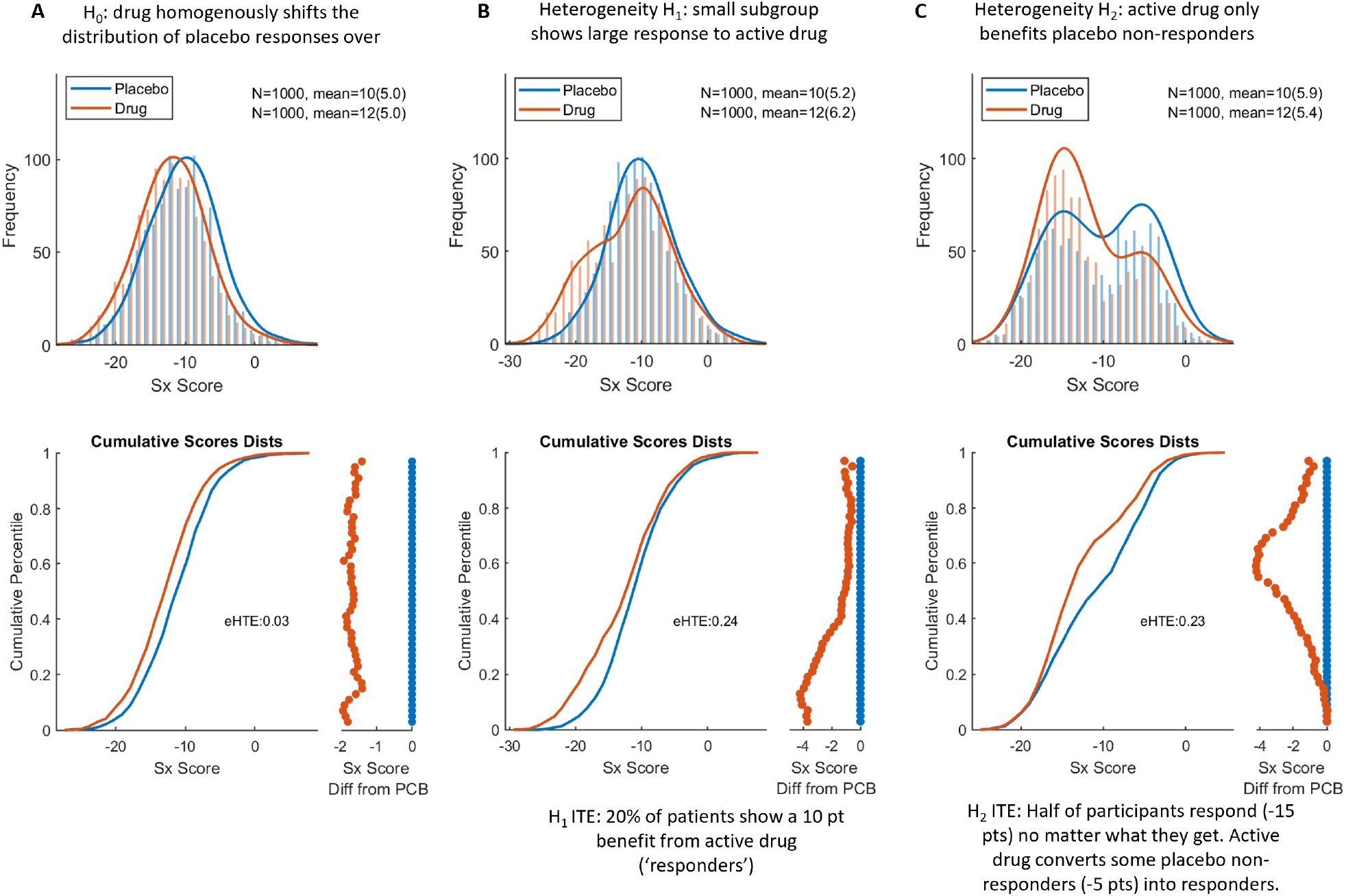
Simulation of treatment heterogeneity. All three simulations include a PCB arm with mean change = -10 and a Drug arm with mean change = -12, with N=1000 participants each. For each simulation, overlapping histograms are shown on top. Sample size, mean, and SD are provided in the inset. Below, cumulative response curves for each arm (matching x-axis). Projected to the right is the difference between the cumulative response curves (matching y-axis). **A)** Simulation 1: The null hypothesis. **B)** Drug responder sub-group (Simulation 2). eHTE = 0.24, P < 0.001. **C)** Placebo responder and non-responder sub-groups (Simulation 3). eHTE = 0.23, P < 0.001.

The computation of eHTE can be expressed mathematically as follows - let *P*(*x*) represent the cumulative response function for the placebo group and *T*(*x*) represent the same for the treatment group, over some range of percentiles *x*∈[*x*1,*x*2,…,*x48*]. For each *xi*, we calculate the difference between the treatment and placebo cumulative responses:

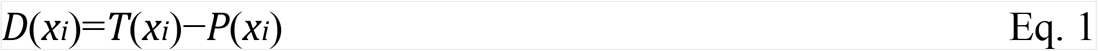

Then, eHTE is calculated as the standard deviation of the differences across percentiles D(xi) divided by the standard deviation of placebo response:

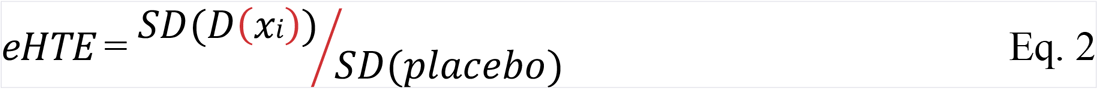

This ratio represents the relative variability in responses between the two arms, normalized by the variability observed within the placebo arm. Importantly, eHTE scales with the standard deviation in the ITE (thus it scales with HTE) and it is unitless (thus it can be compared across different clinical/outcome scales). A high value of eHTE indicates a substantial heterogeneity in responses between the treatment arms (relative to overall variability in placebo response).

### 2.4 Permutation testing and statistical Significance of eHTE

To estimate the probability that observed eHTE values could occur by chance, we simulated normal distributions by randomly selecting the same sample size (as the actual drug/placebo arms) from a normal distribution with the same mean, and standard deviation as the original data. By repeatedly permuting normal placebo and drug distributions (N=1,000 distributions), we generated 1,000 eHTE values under the null hypothesis of no treatment heterogeneity (all individuals respond equally to the treatment). This allowed us to assign a *P*-value, *P*_eHTE_, to the observed eHTE, indicating the likelihood of observing such a value by chance.

### 2.5 Simulation (toy case) Analysis

We simulated three ‘toy cases’ each representing distinct hypotheses regarding treatment response heterogeneity. For each simulations, we set N=1,000 per arm, and pre-defined properties of the placebo and active arm distributions. To demonstrate that eHTE is independent of mean treatment effect, we set mean drug response = -12 and mean placebo response = -10.

1. Null Hypothesis (Simulation A): In this scenario, we simulated a null hypothesis where no heterogeneity of treatment effect exists between the placebo and drug arms. Both arms were generated from normal distributions with standard deviation = 5. ITE = 2 in all participants (SD(ITE)=0).

2. Drug responder sub-group (Simulation B): Here, we simulated a scenario where 20% of individuals in the drug arm experienced a larger improvement (-20 points) compared to the placebo arm, while the remaining 80% experienced improvement that was no different from placebo (-10 points). This scenario aimed to assess the impact of a small subgroup of ‘responders’ in the treatment arms.

3. Placebo responder and non-responder sub-groups (Simulation C): In this scenario, the placebo arm was equally divided into two sub-groups: responders (-15 points) and non-responders (-5 points). The drug arm shifted some individuals from the non-responder group to the responder group, aiming to explore the impact of subgroup responses on overall HTE.

Next, simulation B was used for a power analysis. Specifically, we assessed how large a clinical trial dataset must be to detect HTE; we started with simulation B. Note that this heterogeneous response would appear as a modest treatment effect size = 0.4, and the eHTE is roughly in line with values found in real datasets (Table 1). In this analysis, we generated and tested 28,000 simulations ranging from N=20-300 per arm. For each simulation, a *P*-value was generated by comparing to normal distributions using the permutation approach described above.

**Table 1.** Estimated heterogeneity of treatment effects in psychiatric clinical trials.

| Description | NCTID | Outcome | Arms | eHTE | $P_{\text{eHTE}}$ |
| --- | --- | --- | --- | --- | --- |
| Dasotraline in adults with binge-eating disorder (McElroy et al., 2020) | NCT02564588 | Binge Days Per Week | Dasotraline 4-8mg (159)<br>Placebo (160) | 0.25 | **0.002** |
| Dasotraline in adults with binge-eating disorder: (Grilo et al., 2021) | NCT03107026 | Binge Days Per Week | Dasotraline 4mg (173)<br>**Dasotraline 6mg (150)**<br>**Placebo (163)** | 0.22<br>0.26 | **0.011**<br>0.003 |
| Dasotraline in children with attention deficit disorder (Findling et al., 2019) | NCT02428088 | ADHD RS-IV | Dasotraline 2mg (111)<br>Dasotraline 4mg (115)<br>**Placebo (116)** | 0.14<br>0.16 | 0.49<br>0.32 |
| Dasotraline for the Treatment of ADHD in Adults (Koblan et al., 2015) | NCT01692782 | ADHD RS-IV | Dasotraline 4mg (116)<br>Dasotraline 8mg (115)<br>**Placebo (110)** | 0.14<br>0.19 | 0.48<br>0.17 |
| Dasotraline in adults with ADHD (Adler et al., 2021) | NCT02276209 | ADHD RS-IV | Dasotraline 4mg (219)<br>Dasotraline 6mg (210)<br>**Placebo (219)** | 0.10<br>0.10 | 0.52<br>0.59 |
| Lurasidone or Olanzapine for Schizophrenia (Study D1050231) (Meltzer et al., 2011) | NCT00615433 | PANSS | Luras.40mg (79)<br>Luras.120mg (68)<br>Olanzapine 15mg (87)<br>**Placebo (73)** | 0.14<br>0.23<br>0.21 | 0.79<br>0.18<br>0.20 |
| Lurasidone for acute schizophrenia: a 6-week RCT (Study D1050229) (Nasrallah et al., 2013) | NCT00549718 | PANSS | Lurasidone 40mg (84)<br>Lurasidone 80mg (88)<br>Lurasidone 120mg (86)<br>**Placebo (75)** | 0.12<br>0.21<br>0.22 | 0.83<br>0.19<br>0.15 |
| Lurasidone in the treatment of schizophrenia (Study D1050233) (Loebel et al., 2013) | NCT00790192 | PANSS | Lurasidone 80mg (89)<br>Lurasidone 160mg (96)<br>Quetiapine XR 600mg (99)<br>**Placebo (77)** | 0.15<br>0.16<br>0.19 | 0.50<br>0.37<br>0.19 |
| Psilocybin vs Escital. for Depression (Carhart-Harris et al., 2021) | NCT03429075 | MADRS | Psilocybin (28)<br>**Escitalopram (29)** | 0.36 | 0.153 |
| Psilocybin for Treatment of Major Depressive Disorder (Raison et al., 2023) | NCT03866174 | HAM-D | Psilocybin (50)<br>Niacin (44) | 0.33 | 0.068 |
| Dasotraline or venlafaxine for 8 weeks in adults with MDD (Hopkins et al., 2013) | NCT0058497 | HAMD-17 | Dasotraline 0.5mg (101)<br>Dasotraline 2mg (110)<br>Venlafaxine 150mg (107)<br>Placebo (114) | 0.21<br>0.16<br>0.25 | **0.045**<br>0.24<br>0.034 |

### 2.6 Clinical Trial (participant-level) Analysis

We sought instances for which multiple similar trials were conducted for the same drug/outcome in order to assess if our approach to measuring heterogeneity replicated well. In total, we tested multiple trials of dasotraline for adults with binge eating disorder (Grilo et al., 2021; McElroy et al., 2020), multiple trials of lurasidone (and other antipsychotics) for schizophrenia (Loebel et al., 2013; Meltzer et al., 2011; Nasrallah et al., 2013), multiple trials of dasotraline for attention deficit and hyperactive disorder (Adler et al., 2021; Findling et al., 2019; Koblan et al., 2015), multiple trials of psilocybin for unipolar depression (Carhart-Harris et al., 2021; Raison et al., 2023), a trial of non-racemic amisulpride for BPAD depression (Loebel et al., 2022), and a trial of Venlafaxine for MDD (Hopkins et al., 2013). Details are provided in Table 1.

## 3 Results

### 3.1 Simulation of treatment heterogeneity

#### 3.1.1 Figure 1

To gain intuition on how different types of response subgroups might impact eHTE (and the associated *P*-values and power), we simulated three ‘toy’ cases. Each had a 2 point benefit of drug over placebo and a realistic effect size around ∼0.4. In the null hypothesis case, drug has equal benefit on all participants (HTE=0) (Figure 1A). Two alternative cases present intuitive ways in which H_o_ can be rejected (independently from mean treatment effect size): B) a small subgroup of patients show large response to active drug (while the rest do not separate from placebo), and C) a subgroup responds to drug or placebo, a second subgroup responds to active drug but not placebo, and a third subgroup does not respond to either (Figure 1B-C). For simulations B and C, eHTE was larger than all permutations of normal distributions (P < 0.0001). Note that in H2, the standard deviation is smaller in drug arm than placebo arm, indicating that the ‘variability ratio’ approach would assert that heterogeneity is absent.

It may help to imagine our approach like this – you have 100 individuals in the placebo arm, and 100 individuals in the drug arm - you line each group up in order of clinical response and then pair them. Then compare the 3^rd^ largest drug responder, to the 3^rd^ largest placebo responder, the 5^nd^ largest drug responder, to the 5^nd^ largest placebo responder, and so on down the line comparing pairs. These comparisons are shown by the orange dots on the bottom right of each figure. eHTE is the horizonal deviation of the orange dots. A vertical line ‘|’ of orange dots (e.g. Fig 1A, bottom) is an eHTE of 0. Alternatively, a subgroup of responders to active drug (H1) could cause a large drug-placebo separation in the top percentiles of responders, resulting in a non-vertical (‘/’ shaped) drug-placebo difference across percentile. Alternatively, a drug that only benefits placebo non-responders (H2) would cause a drug-placebo separation only in the lower percentiles of responders, resulting in a nonvertical (‘\’ or ‘<’ or some other shape) drug-placebo difference.

### 3.2 Power to detect subgroups

Next, we used simulated cases for a power analysis. Specifically, we asked how big a clinical trial dataset must be detect a fairly large HTE; we started with simulation B - 20% of the population has an effect size = 2 (ITE=2*SD_placebo_), and 80% show no difference from placebo (ITE=0). With the eHTE approach, a total sample size of N=200 (or N=100 each for drug and placebo arms) would have ∼80% power to detect significant (α = 0.05) heterogeneity (Fig. 2). Thus, our approach should have power to identify HTE in most Phase 3 studies in psychiatry, should HTE exist.

**Figure 2.**
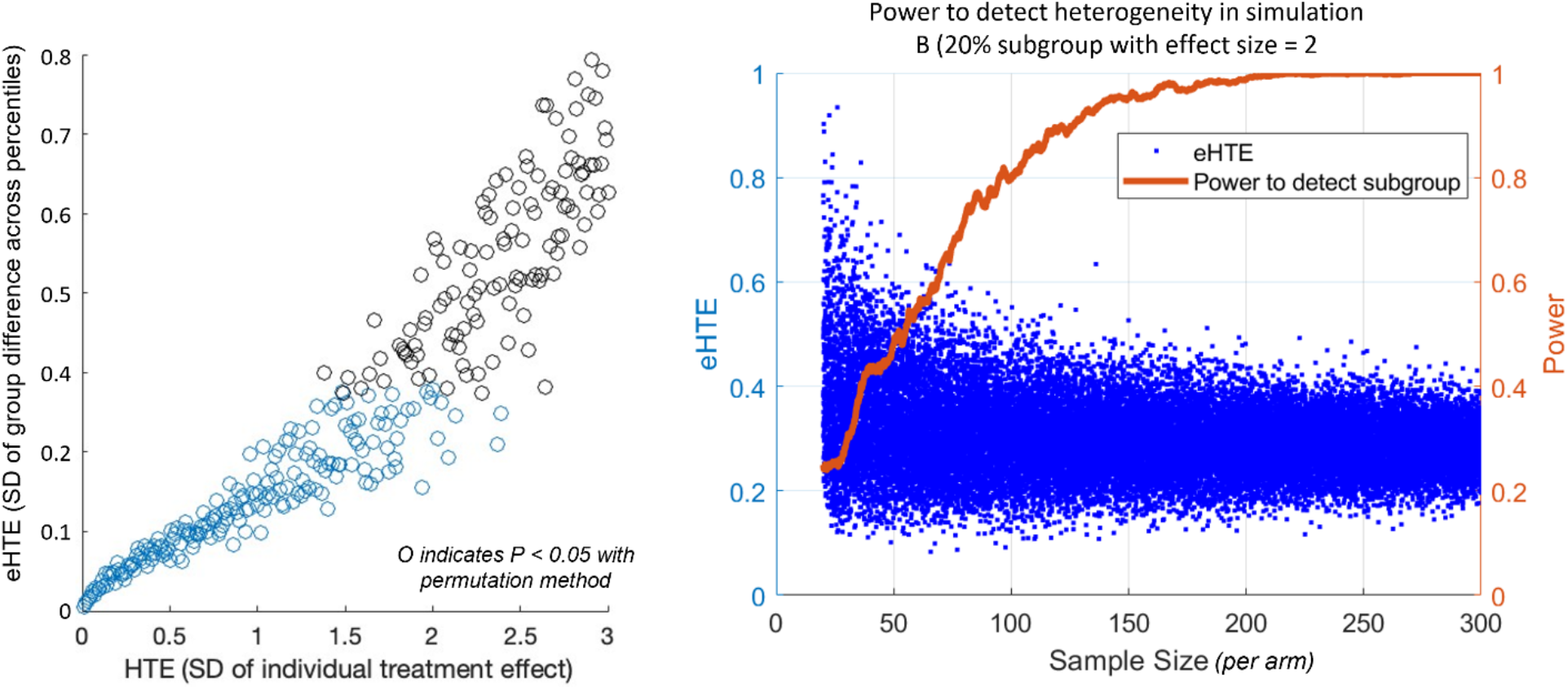
Power to detect treatment heterogeneity with eHTE. Left: A comparison between eHTE and actual HTE (computed by gradually increasing the standard deviation of the individual treatment effect in Simulation A). X-axis untis are scale dependent and therefore arbitrary. **Right:** Based on simulation 1B – 20% of the population has an effect size = 2, 80% show no difference from placebo). Blue dots represent eHTE at across a range of sample sizes. Orange line depicts power to detect a significant heterogeneity with alpha set to p<0.05.

#### 3.2.1 Figure 2

### 3.3 Participant-levels datasets

We tested this method for visualizing and quantifying heterogeneity of treatment effect in 11 psychiatric clinical trial datasets with 23 active treatment arms (Table 1) for which participant-level outcomes were available (note that most meta-analyses rely on databases which include group statistics but not subject-level data). Multiple treatments showed significant heterogeneity of treatment effect (*P*_eHTE_ < 0.05 uncorrected).

#### 3.3.1 Examples of different types of heterogeneity in Real Data

Visualizing participant-level outcomes data revealed important intuitions about the nature of drug responses which are obscured by group-level statistics and figures (Fig. 3). Importantly, these qualitative instances of non-normal and non-homogenous treatment effects were well captured and quantified by the eHTE approach. Below, we describe some cases in which treatment heterogeneity was or was not reliably detected despite drug-placebo separation.

**Figure 3.**
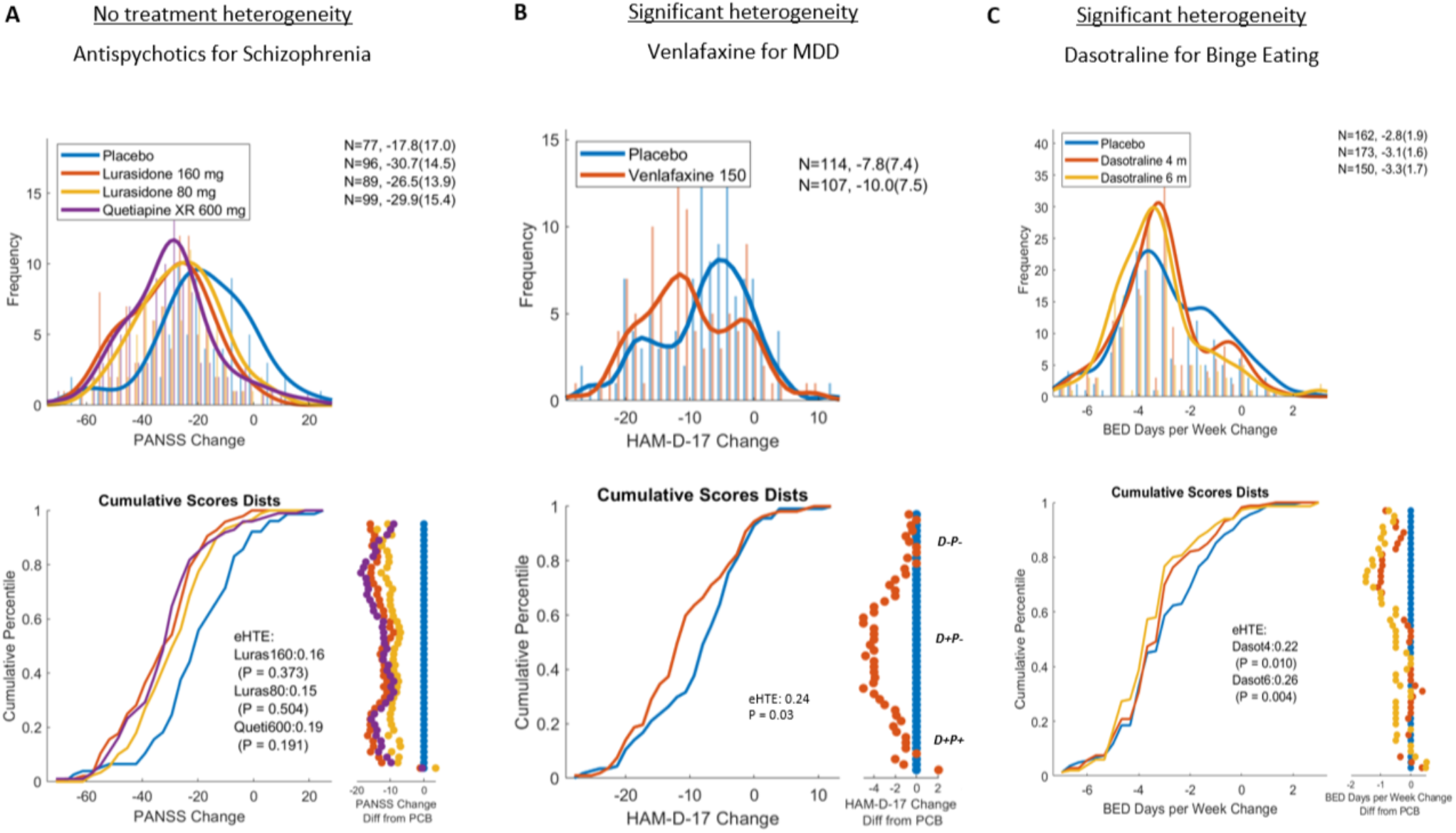
Heterogeneity in published clinical trials. Three examples from Table 1 are visualized. A). Lurasidone and Quetiapine in the treatment of schizophrenia (Study D1050233) (Loebel et al., 2013) **B)** Venlafaxine vs placebo in the treatment of major depressive disorder **(Hopkins et al., 2013) C)** Phase 3 RCT of dasotraline in adults with Binge eating disorder (Grilo et al., 2021).

A Phase 3 randomized controlled trial (RCT) of Lurasidone and Quetiapine versus placebo for schizophrenia (Loebel et al., 2013) revealed no heterogeneity of treatment effect (*P*_eHTE_ > 0.1) in all active arms. A lack of heterogeneity in drug response replicated across all arms of 2 additional RCTs (with 6 active arms) of antipsychotic for schizophrenia (Table 1), supporting the validity of this negative observation.

A clinical trial of Venlafaxine for major depressive disorder revealed significant heterogeneity of treatment effect (Fig 3B; *P*_eHTE_ = 0.034). Similar to the simulation C above, a subgroup in both arms showed a large response, a middle subgroup showed a moderate response that separated drug from placebo, and yet another subgroup showed no response on drug or placebo.

A Phase 2 trial of Dasotraline for treatment of binge eating disorder (BED) (McElroy et al., 2020) showed significant heterogeneity (*P*_eHTE_ = 0.002) – the placebo arm appeared bimodal with placebo responder and non-responder subgroups – and active treatment only benefited placebo non-responders. Supporting the validity of this observation, a Phase 3 trial of Dasotraline for BED (Grilo et al., 2021) showed exactly the same type of heterogenous response at both doses (Fig. 3C; *P*_eHTE_ = 0.011 for 4mg dose, *P*_eHTE_ = 0.003 for 6mg dose).

It is notable that for the trial of Venlafaxine for MDD trial and the trials of Dasotraline for Binge Eating disorder, eHTE was significant despite the standard deviation of responses being nearly equal between placebo and active arms (i.e. variability ratio was close to 1).

## 4 Discussion

We described a novel analytical index, eHTE, to quantify heterogeneity of treatment effects in clinical trial datasets. We demonstrated, through simulations, that eHTE can be used to go from intuition - how subpopulations may differ in response to a treatment – to quantitative measurements and hypothesis testing. Most importantly, we computed eHTE in multiple clinical trial datasets (with participant-level data) and found that heterogeneity replicated across studies testing the same drug for the same indication (e.g., Dasotraline for binge eating disorder). In instances in which significant heterogeneity was detected, the specific ‘shape’ of individual treatment effects varied – with some instances of heterogeneity matching our a priori hypotheses (Fig. 1) well, and others not so clearly fitting a priori simulations. Taken together, these observations support the assertions that A) treatment heterogeneity is present in psychiatric clinical trials, and B) the eHTE approach provides a stable metric of heterogeneity.

Primary outcomes in clinical trials are typically first-order statistics, relating to central tendencies of the drug and placebo arms. On the other hand, eHTE deals with second-order statistics, pertaining to the variability of responses in each arm. Because eHTE provides insights into the variability of treatment effects independent of central tendency, it offers a distinct perspective. This means that reporting heterogeneity using eHTE may not necessitate multiple comparisons correction of efficacy-related statistics. Therefore, eHTE can offer a more nuanced understanding of treatment effects without additional statistical cost.

Conceptually, the approach of comparing across response percentiles (which can be thought of as matching up ‘pairs’ of individuals in treatment and placebo group) is a useful way to approximate individual treatment effects (the hypothetical difference between a person’s outcome on treatment drug vs placebo). It can detect significant heterogeneity in intuitive instances – such as when a subgroup of patients respond to drug and the rest do not. And it can reliably do so at sample sizes typically used in clinical trials in psychiatry and neurology (Fig. 2).

A previously proposed method to assess heterogeneity using ‘variability ratio’ found no evidence of heterogeneity in antidepressant response (Maslej et al., 2021; Nakagawa et al., 2015; Volkmann et al., 2020). This approach assumes that if HTE is present, then variability should be larger in drug arm than placebo arm (because variability of placebo effect and drug effect (ITE) are additive in the drug arm). Crucially, this approach relies on the assumption of no interaction between placebo response and drug effect (ITE). In multiple datasets, we found evidence of heterogeneity despite no difference in standard deviation between arms (Fig. 3 – Venlafaxine for MDD, Dasotraline for BED). This strongly suggests that the variability ratio method is flawed. Indeed a negative interaction between placebo response and ITE is supported by meta-analyses observing that larger placebo response correlates with smaller drug-placebo separation in depression (Iovieno & Papakostas, 2012) and schizophrenia (McCutcheon et al., 2022).

BOX: Relevance of treatment heterogeneity to Enrichment

In “Enrichment Strategies for Clinical Trials to Support Determination of Effectiveness of Human Drugs and Biological Products: Guidance for Industry” the Food and Drug Administration encourages drug developers to consider three enrichment strategies ***(Center for Drug Evaluation and Research, 2019)***:

1. Strategies to decrease variability (e.g. ‘excluding patients whose symptoms improve spontaneously’)

2. Prognostic Enrichment (identify patients with high risk of disease-related endpoint)

3. Predictive Enrichment (identify responders to active drug)

The measurement of treatment heterogeneity has direct implications on when to employ these enrichment strategies in large clinical trials. In comparing the distribution of treatment effect between study arms, distinct patterns of heterogeneity may emerge. For example, you may observe a subgroup of individuals on active drug show complete remission (as in simulation B), or a subgroup of individuals show remission in both arms (‘D+P+’), but active drug decreases the number of non-responders (‘D+P-’) (as in simulation C). Below, we diagram how these patterns can be used to inform enrichment strategies in subsequent trials.

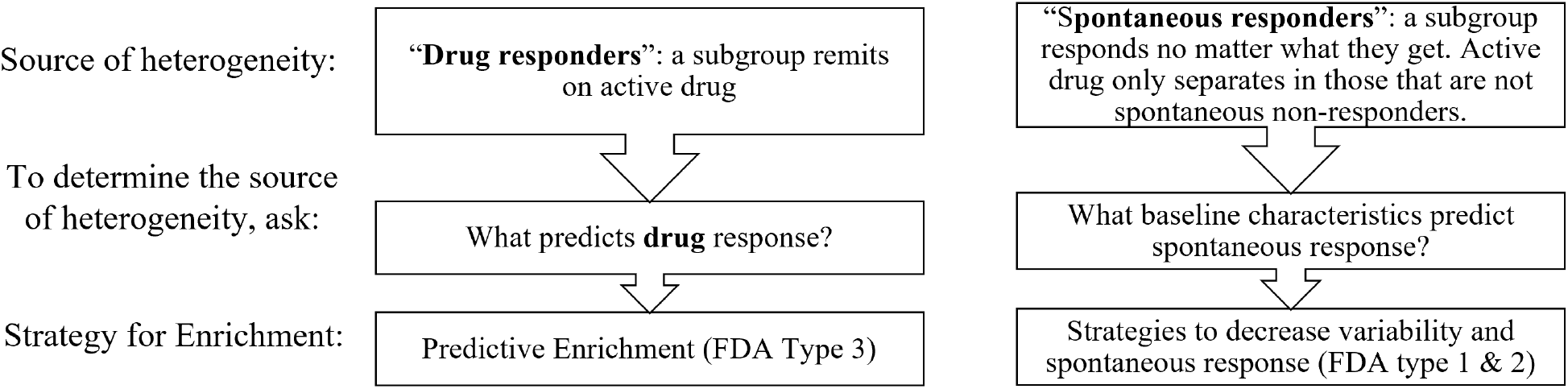

Because meta-analyses have classically reported the mean treatment effect or effect size, the majority of clinical trial databases do not provide subject-level data that would be needed to examine heterogeneity. To realize the utility of eHTE and other approaches to understanding heterogeneity and subgroup analyses, it is imperative that subject level data from clinical trials be made publicly available.

### 4.1 Antidepressant Subgroups (D+P+, D+P-,D-P-)

Previous explorations of depression clinical trials have described 3 groups based on response to treatment (Fava, 2003, 2015). The “D−P−” population comprises patients who are not responsive to active treatment (D) and placebo treatment (P). The “D+P+” population comprises patients who are responsive to either active (D) or placebo (P) treatments. The “D+P-” population, those who response to active but not placebo, are the entire source of individual treatment effects and drug-placebo separation. This description fits well with our observation. In many of the trials with significant heterogeneity, we observed a subgroup in both arms showed a large response, a middle subgroup showed a moderate response that separated drug from placebo, and a third subgroup showed no response on drug or placebo.

### 4.2 Personalized medicine

Measuring and reporting eHTE will allow for better clinical guidance. Based on standard efficacy outcomes, a provider might tell their patient with depression “Venlafaxine shows a slight (2 points on HAM-D) benefit compared to placebo”. But using eHTE (e.g. Fig 3B), the provider could instead say, “half of patients who start Venlafaxine show a substantial benefit (4 points on HAM-D) compared to placebo and half of patients don’t show any benefit beyond placebo.” This provides more accurate and useful prognostic information.

### 4.3 Connecting eHTE to covariates/subgroups

The measurement of treatment heterogeneity provides an empirical basis for deciding when and where we might hypothesize the presence of subgroups. As described in Box 1, the existence of significant HTE is a useful prior in testing if specific baseline characteristics differentiate drug (or placebo) responders from non-responders. In a circumstance where a specific baseline characteristic is causing treatment heterogeneity, eHTE measured in homogenous subgroups should be less than eHTE in the full heterogeneous cohort.

eHTE similarly may provide insight into what specific type of enrichment strategy would be warranted (Box 1). For example, a scenario where you observe heterogeneity due to subgroup of responders to active drug (i.e., Fig 1B and Fig 3A) would motivate a predictive enrichment strategy focused on subgroup selection (FDA type 3). Whereas a subgroup that responds equally to placebo or drug would motivate an enrichment strategy to remove placebo responders (FDA type 1 or 2).

### 4.4 Conclusion

The eHTE approach offers nuanced insights into treatment heterogeneity within clinical trials, moving beyond conventional mean comparisons and allowing for more refined interpretations of trial data. It eases the identification of differential response patterns between trial arms, thus holding significant implications for not only for prospective clinical trial enrichment, but also for personalized medicine and the development of targeted therapeutic strategies. There is nothing about this approach that is specific to psychiatry. There may be utility in measuring heterogeneity of treatment effect across any area of medicine.

## Data Availability

Code to compute eHTE on participant-level clinical data is available in Python, Matlab, and SAS languages at http://gitlab.com/siegelandthebrain. Some of the clinical trial data are proprietary to Sumitomo Pharma America and are available upon request.

## Code availability

Code to compute eHTE on participant-level clinical data is available in Python, Matlab, and SAS languages at http://gitlab.com/siegelandthebrain.

## Declaration of generative AI and AI-assisted technologies in the writing process

During the preparation of this work the author(s) used OpenAI ChatGPT in order to develop and optimize some analysis code. After using this tool/service, the author(s) reviewed and edited the content as needed and take(s) full responsibility for the content of the publication.

## Disclosures

Authors JSS JZ ST AO STS KSK and SCH are employees of Sumitomo Pharma America. In the past year, JSS has received consulting fees from Forbes Manhattan and Longitude Capital. In the past year, Dr. Faraone received income, potential income, travel expenses continuing education support and/or research support from Aardvark, Aardwolf, AIMH, Tris, Otsuka, Ironshore, Johnson & Johnson/Kenvue, ADHDOnline, KemPharm/Corium, Akili, Supernus, Atentiv, Noven, Sky Therapeutics, Axsome and Genomind. With his institution, he has US patent US20130217707 A1 for the use of sodium-hydrogen exchange inhibitors in the treatment of ADHD. He also receives royalties from books published by Guilford Press: Straight Talk about Your Child’s Mental Health, Oxford University Press: Schizophrenia: The Facts and Elsevier: ADHD: Non-Pharmacologic Interventions. He is Program Director of www.ADHDEvidence.org and www.ADHDinAdults.com. Dr. Faraone’s research and education programs are supported by the European Union’s Horizon 2020 research and innovation programme under grant agreement 965381; NIH/NIMH grants U01AR076092-01A1, R0MH116037, 5R01AG064955-02, 1R21MH126494-01, 1R01NS128535-01, R01MH131685-01, 1R01MH130899-01A1, Corium Pharmaceuticals, Tris Pharmaceuticals and Supernus Pharmaceutical Company.

## Notes

### Competing Interest Statement

Authors JSS JZ ST AO STS KSK and SCH are former employees of Sumitomo Pharma America. In the past year, JSS has received consulting fees from Forbes Manhattan and Longitude Capital. In the past year, Dr. Faraone received income, potential income, travel expenses continuing education support and/or research support from Aardvark, Aardwolf, AIMH, Tris, Otsuka, Ironshore, Johnson & Johnson/Kenvue, ADHDOnline, KemPharm/Corium, Akili, Supernus, Atentiv, Noven, Sky Therapeutics, Axsome and Genomind. With his institution, he has US patent US20130217707 A1 for the use of sodium-hydrogen exchange inhibitors in the treatment of ADHD. He also receives royalties from books published by Guilford Press: Straight Talk about Your Child?s Mental Health, Oxford University Press: Schizophrenia: The Facts and Elsevier: ADHD: Non-Pharmacologic Interventions. He is Program Director of www.ADHDEvidence.org and www.ADHDinAdults.com. Dr. Faraone's research and education programs are supported by the European Union?s Horizon 2020 research and innovation programme under grant agreement 965381 NIH/NIMH grants U01AR076092-01A1, R0MH116037, 5R01AG064955-02, 1R21MH126494-01, 1R01NS128535-01, R01MH131685-01, 1R01MH130899- 01A1, Corium Pharmaceuticals, Tris Pharmaceuticals and Supernus Pharmaceutical Company.

### Clinical Trial

Data are used from: NCT02564588 NCT03107026 NCT02428088 NCT01692782 NCT02276209 NCT00615433 NCT00549718 NCT00790192 NCT03429075 NCT03866174 NCT0058497

### Funding Statement

Yes

### Author Declarations

This is a meta-analysis. No data were collected as a part of this research.

